# Modelling the Dynamics of SARS-CoV-2 during early phase of infection

**DOI:** 10.1101/2025.02.12.25321951

**Authors:** Jingsi Xu, Martín López-García, Thomas House, Ian Hall

## Abstract

Interpreting viral mechanism of SARS-CoV-2 based on human body level is critical for developing more efficient interventions. Due to the limitation of data, limited models consider the viral dynamics of early phase of infection. The Human Challenge Study Killingley et al. (2022) enables us to garner data from the inoculation to the 14th day after the infection, which provides an overview of the SARS-CoV-2 within host infection dynamics. In the Human Challenge Study, each volunteer was inoculated with 10TCID50, approximately 55PFU, of a wild type of virus (Killingley et al. (2022)), and the data indicates that the viral load reduced below the detectable level within a day.

The simplified within host models developed by Xu et al. (2023) explain the data from the Human Challenge Study (Killingley et al. (2022)). However, they do not explain the viral decay from Day 0 to Day 1. Hence, in this paper, we aim to develop a new viral mechanism to explain this phenomenon. Based on the simplified within host models developed by Xu et al. (2023), we consider that the virus will first go through an adjustment phase and then start to replicate. A new dose-response model is developed to evaluate the probability of infection by constructing a boundary problem. We will discuss this viral mechanism and fit the model to the data of the Human Challenge Study (Killingley et al. (2022)) by adopting AMC-SMC (approximate Bayesian computation-sequential Monte Carlo). Based on the results of parameter inference, we estimate that the adjusted viral load is around 1% of the inoculated viral load.

## 1. Introduction

Since the pandemic of SARS-CoV-2, many studies report different within-host models aiming to introduce and explain different viral mechanisms (see e.g. Challenger et al. (2022), Goyal et al. (2020), Gonçalves et al. (2021),Li et al. (2022), Ghosh (2021),Sadria & Layton (2021),Du & Yuan (2020), Hernandez-Vargas & Velasco-Hernandez (2020),Abuin et al. (2020),Li et al. (2020), and Wang et al. (2020)). Understanding different models of viral mechanisms may offer more insights into considering targeted and effective intervention to limit the spread of SARS-CoV-2, and other respiratory diseases.

The Human Challenge Study (Killingley et al. (2022)), abbreviated hereafter to HCS, measured the detectable viral load in the upper respiratory tract. In the study, 34 young male volunteers who were between 18-29 years old and had not been previously vaccinated or infected were given 10TCID50 of a wild type of virus, where 10TCID50 is approximately 55PFU with credible interval (Killingley et al. (2022)). A few days after the inoculation, the viral load is observed to drop below the detectable level, 5PFU, subsequently growing above the detectable level. The length of days under detectable level thus varies from mid-turbinate to throat. The throat data shows a shorter period ranging from 1 day to 3 days with a mean of 2.5 days while the mid-turbinate ranges between 2 days to 8 days with a mean of 3.7 days. Previous work developed a simplified within-host model, Xu et al. (2023), assuming two mechanisms for the decay of viral load, 1) by the depletion of susceptible cells and 2) the adaptive immune response respectively, both of which explain the HCS data well separately and in combination. However, this model does not explain the early viral decay and so does not use the initial dose from HCS. In this paper, we investigate a viral mechanism to explain the earlier low viral load measures and extend the models of Xu et al. (2023) to consider an adaptive immune response growing logistically rather then a fast switch.

As we are using data from HCS rather than seek to model onset of symptoms (or infectiousness) explicitly (i.e. modelling the incubation or latent periods) we define early infection as the time before detectable viral load measured. It has been reasonably suggested that an ‘eclipse phase’ may be added into the SIV model system to help explain the early decay of the viral load, in which the susceptible cells first become infected cells in eclipse phase and then switch to productively infected cells (cf. Wang et al. (2020)). There are many studies that consider the eclipse phase (cf. Hernandez-Vargas & Velasco-Hernandez (2020),Beauchemin et al. (2008), Holder et al. (2011), Madelain et al. (2018), Williams et al. (2024) and Baccam et al. (2006)). This may be a biological reality but is likely on a timescale faster than the cadence of data collection from cases recruited to studies.

Different from the concept of ‘eclipse phase’, our core assumption here is that during the first few days after the exposure, there is a period of viral ‘adjustment’. Specifically, we assume that there exists a relatively short period after the inoculation, during which the viral load will only decay. After this period, the virus will have adjusted to human body and start to replicate. There are then two processes that may cause an individual to not become infected upon exposure. Firstly the given exposed viral load may decay to extinction during the adjustment phase. Secondly the residual viral load will have a chance of ‘fading out’ during the replication phase prior to ‘infection’. Therefore, we can develop a dose-response model to estimate the chance of infection.

The viral load of SARS-CoV-2 has been linked to the degree of disease severity, infectiousness, lung damage and transmission risk (see e.g. Fajnzylber et al. (2020), Williamson et al. (2020) and Pujadas et al. (2020)) though the HCS found limited evidence of viral load corresponding to more sever infection (Killingley et al. (2022)). The magnitude of the viral load is then an important determinant to evaluate transmission rate,(Watanabe et al. (2010)), and patients with higher viral load can be closely epidemiologically related (Marks et al. (2021)). The Hill function has been widely adopted to link viral load and transmission risk for different diseases including influenza (cf. Handel & Rohani (2015)) and SARS-CoV-2 (cf. Heitzman-Breen & Ciupe (2022),Ke et al. (2020), and Goyal et al. (2021)). The transmission risk is split into contagiousness and infectiousness in Goyal et al. (2021), which is evaluated by the Hill function with same parameters. In Ke et al. (2020), the probability of infection is defined as the chance that at least one virion seeds infection and they assume only a proportion of inhaled virus could arrive the respiratory tract of the contact. Based on these assumptions, an exponential form dose-response model with a Michealis-Menten term representing the amount of virus shed from the upper respiratory tract is adopted in Ke et al. (2020). For the dose-response model developed by Ke et al. (2021), it can be simplified to a Hill function to link the viral load with the infectiousness if the composite parameter is sufficiently small. Furthermore, Ke et al. (2021) indicates that the infectiousness is sub-linear with the viral load and suggests that logarithm of viral load is a better surrogate of infectiousness when data is given by RNA copies. Moreover, Haas et al. (2014) introduces a series of dose-response models under the competing risk framework for bacterial infections, and two models, the exponential and approximate beta-Poisson dose-response models, of Haas et al. (2014) are applied in Xu et al. (2023) to consider the transmission risk of SARS-CoV-2. Ejima et al. (2021) estimate the infection establishment threshold according to each patient, however, it is measured by RNA copies, which is not applicable in our case.

In this work, we also aim to develop an alternative derivation for a dose-response model. The simplified model developed in Xu et al. (2023) is adapted to reduce the viral dynamics to a stochastic differential equation (SDE) that only involves the virus state, from which we construct a boundary problem to evaluate the probability of infection. To our knowledge the boundary problem approach has not been used in deriving dose-response models.

This study then involves three extensions to the model proposed in Xu et al. (2023): 1) We consider early infection dynamics, 2) merge this with the stochastic fade out properties of the system to use evidence of infecting dose in model calibration, and 3) we extend the model used in the replication phase to include a smoothly increasing adaptive immune response.

## 2. Methods

Consider a contact who inhales a certain amount of virus during an exposure event. Whether this contact will develop an infection depends on the viral dynamics during the early phase of infection. Specifically, we define the early phase infection as the time between the inhalation and the time that virus is above the detectable level in those cases that develop infection. During this early phase of infection, there are two independent events that may happen: viral adjustment and then replication. During the viral adjustment phase the inhaled dose reduces so that only a portion of the virus remains in the host. After some time the residual virus successfully enters susceptible cells, starts to replicate and if the viral load reaches the threshold for infection, the contact will develop an infection.

### 2.1. Viral adjustment phase

After inhalation, we assume that the virus will decay during the viral adjustment phase and only a small amount of the virus will achieve viral colonisation. In this case, identifying the amount of virus that survives is important to estimate the probability of subsequent infection. This is conceptually similar to the rapid deposition phase expected in bacterial infections (Heppell et al. (2017)) but here deposition is only a single component of the adjustment process.

To simulate this parsimoniously, we assume that the virus will decay exponentially with respect to time:

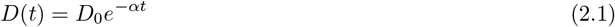

for *t* ∈ ℝ_+_. In (2.1), parameter *α >* 0 measures the viral decay rate during the viral adjustment phase and *D*_0_ is the initial dose the contact inhaled during the exposure. This model suggests that after inhalation, the virus will be exhaled during breathing or cleared by immune response, and at *t* = *t*\*, a certain amount of virus, 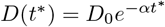, realises viral colonisation at the location of the infection.

A deterministic model would always predict some residual viral load that would cause infection after the switch to the replication phase (cf. the ‘atto-fox’ phenomena (Mollison (1991))). Instead, we develop a stochastic version where the dynamics during the viral adjustment phase is given by:

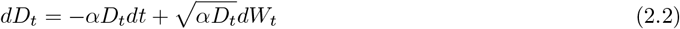

for *t* ∈ [0, *t*\*] where *t*\* is the time the virus takes to get adjusted to human body and the diffusion term allows *D*_*t*_ to hit 0. In this case, it is very important to understand the probability of clearance during the viral adjustment and what is the distribution of the viral load that successfully entered the susceptible cells. Due to the structure of (2.2), there is no analytical form of the probability of *D*_*t*_ hitting 0 before *t*\*, as the PDE of the corresponding boundary problem or time-space method (see cf. Pedersen & Peskir (2016)) has no analytical solution. Hence, we will carry out simulation to evaluate the viral shedding under the stochastic setting in the following analysis. Instead of the stochastic diffusion model in (2.2) a discrete event stochastic simulation could be developed. This was considered, see Supplementary material, but the results were not very different to the diffusion-based simulation. Moreover, we note that a unit value of PFU is not a single viable entity and so fractional amounts of PFU have biological meaning.

### 2.2. Viral replication phase

The simplified within-host model developed in Xu et al. (2023) provides a viral mechanism when viral load is above the detectable level, which is given by:

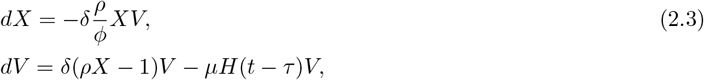

where*H*(*t* − *τ* ) is a Heaviside function and *τ* is the time when the adaptive immunity response is activated. In (2.3), X represents the compartment of susceptible cells, while V represents the compartment of viral load. Furthermore, by the definition of Xu et al. (2023), the initial condition of *X* and *V* are *X*(0) = 1 and *V* (0) = *V*_0_, and *ρ* represents the threshold parameter in the model (so deterministically if *ρ >* 1 the virus grows and if *ρ <* 1 it decays) whilst *ϕ* modifies the removal of susceptible cells. One of the advantages of this model is that it parsimonious, however, it is still not possible to distinguish between depletion of susceptible cells and the increasing adaptive immune response based on observational data like that provided by the HCS alone.

We can relax the assumption in (2.3) that the adaptive immune response is a Heaviside step function so that the immune response grows logistically over some timescale *T* which controls the speed of immune response growth. We also recast the model parameters for future calibration. Defining *ρ* = 1 + *θ/ν, δ* = *νσ/T* and *µ* = *σ/T* for some parameters *σ, θ* and *ν* means we have a model

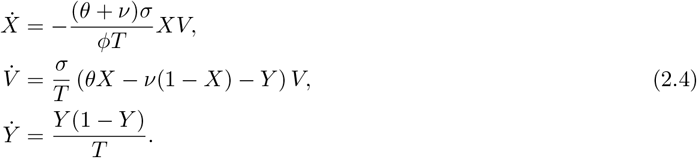

Note that the role of parameters *σ, θ*, and *ν* is to reparameterise the model, which has no biological meaning. The variable *Y* then is a non-dimensional representation of immune response scaling from a small contribution at initial time *t* = 0 to 1 as *t* → ∞. Notice that when *X* ∼ 1 and *Y* ∼ 0 this system will grow exponentially in *V* with rate *r* = *θσ/T* , and when *X*∼ 1 and *Y*∼ 1 it will decay exponentially with rate *r*_*D*_ = (1 − *θ*)*σ/T* . Then introducing a time *τ* we may solve the adaptive immune response directly such that *Y*_*t*_ = *θ*(*θ*+(1− *θ*)*e*^−(*t*−*τ*)*/T*^ )^−1^ which means that at time *τ* the adaptive response has achieved *θ* of its eventual impact which also means that the parameter *θ* ∈ [0, 1].

We could solve this in the composite situation of both susceptible cell depletion and immune response increase. However as noted in Xu et al. (2023) with (2.3) the contributing mechanism is unidentifiable in this scenario given the data provided from HCS. If we limit consideration to the case where *X* ∼ 1 (so assume that *ϕ* ≫ *V* (*t*) for all *t*) then *τ* is the time of peak viral load (noting that when *t* = *τ* we have 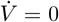). The parameter *τ* then can be considered identical to the *τ* that appears in the Heaviside function in (2.3). We can define a time *t*_*L*_ representing the time at which the viral load is above some (known) detectable level (*V*_*L*_) for the first time, meaning that

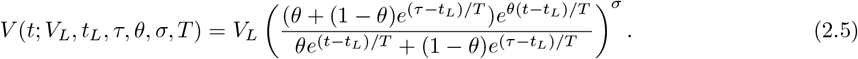

### 2.3. Probability of infection and boundary problems

To find the probability of infection during this replication phase, we introduce stochasticity into the viral dynamics in Equation (2.4) to describe the heterogeneity of response across patients. Hence, the viral dynamics, *V*, after the viral adjustment is defined by the following SDE (assuming *Y* = *Y* (*t*\*) and *X* ∼ 1):

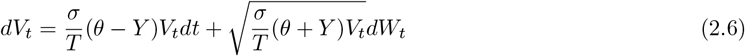

for *t* ∈ [*t*\*,∞) and 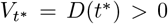 is the amount of virus deposited at time *t*\*. Furthermore, we set that E_*t*,*v*_(*V*_*t*_) *<*∞ for any (*t, v*) ∈ [*t*\*, ∞) × R. After the virus deposited at *t*\* *>* 0, the contact will be infected if the viral load eventuallyreaches the threshold, *M >* 0, for infection, i.e. *V*_*t*_ = *M* for some *t > t*\*, or the contact will clear the virus before it approaches to the threshold for infection, i.e. *V*_*t*_ = 0 for some *t > t*\*. Whilst we set *M* = *V*_*L*_ the limit of detection we do not have to make this restriction but simply note that *M > V* (*t*\*) for the rest of the paper to avoid regularity issue. The goal of this section is to find the relationship between the adjusted viral load, *D*(*t*\*), and probability of infection *P*_*I*_ (*D*_0_) for a given dose *D*_0_. Note that 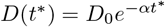 so we can link the probability of infection with initial dose *D*_0_. During the viral replication phase, there are only two events will happen, infection and clearance, so we can define two stopping times:

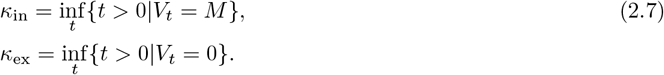

Finding the probability of infection is equivalent to calculating the probability of P(*κ*_in_ *< κ*_ex_). To estimate this probability, we first need to obtain the expectation 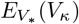 as 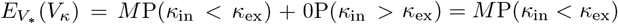, where *κ* = min{*κ*_*in*_, *κ*_*ex*_}.

For SDE (2.6), we see that the drift term satisfies (*σ*(*θ* − *Y* )*V/T* )^2^ = (*θ* − *Y* )^2^(*σV* )^2^*/T* ^2^ ≤ *K*(1 + |*V* |^2^), and the diffusion term satisfies 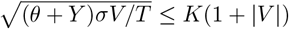 for some constant *K >* 0 (remember that *Y < θ* for *t < τ* so this condition will hold for times prior to the peak viral load). Based on the continuity and strict positivity of (*θ* − *Y* )^2^*V* ^2^, the SDE (2.6) has an unique strong solution with strong markov property (cf. Stroock-Varadhan Theorem on Chapter V of Rogers & Williams (2000)). Define a value function *F* (*v*) = *E*_*v*_(*V*_*κ*_), we can form the following boundary problem with infinite time horizon (cf. chapter 3 of Peskir & Shiryaev (2006)), which is given by:

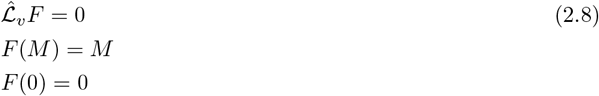

in which 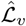 is the infinitestimal generator and *F* (*M* ) = *M* and *F* (0) = 0 describe the corresponding boundary conditions. Solving Equation (2.8) will give *E*_*v*_(*V*_*κ*_), hence, recalling 𝕃_*v*_*F* = 0, one has:

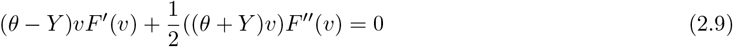

(noting the factor *σ/T* drops out). Note that 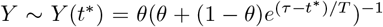. Solving Equation (2.9) gives:

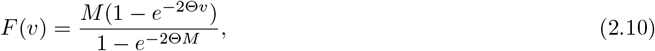

where:

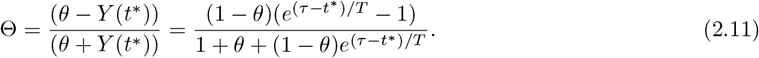

Since we have known that *E*_*v*_(*V*_*κ*_) = *M* P(*κ*_in_ *< κ*_ex_), the probability of infection during the replication phase can be easily obtained:

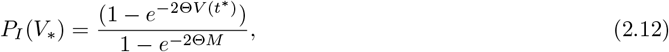

where *V* (*t*\*) is the amount of residual virus following the adjustment phase. In Equation (2.12), it is reasonable to assume that *M* varies for different patients and obtaining the actually value for *M* can be challenging, which leads to unidentifiablity for parameter inference. Therefore, we would like to investigate what happens when *M* tends to infinity. Hence, when *M* → ∞, the probability of infection is:

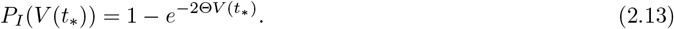

Note that (2.13) has the same form as the exponential dose response function. However, since 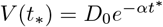, Equation (2.13) gives:

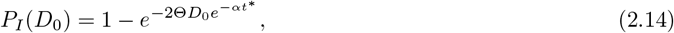

which links the initial dose *D*_0_ and the probability of infection with parameters arising from the model calibration to HCS data, though given an initial dose this is still of the form of the exponential dose-response function.

### 2.4. Parameter inference

As mentioned in introduction, we will use the data from the HCS (cf. Killingley et al. (2022)). The data collected from this study involving throat and mid-turbinate swabs from 18 volunteers for the first 14 days after infection and was recorded by culture and qRNA. Since we only consider the potential of transmission risk in this paper, we will only use the culture data in the following part. First the replication phase model (2.5) is calibrated to the individual trajectories (with *V*_*L*_ = 5 PFU fixed) to identify parameters *t*_*L*_, *T* , *σ, τ* and *θ*.

We perform approximate Bayesian computation - sequential Monte Carlo (ABC-SMC) in which a multivariate normal distribution with optimal local covariance matrix is adopted to pursue parameter inference (cf. Toni et al. (2009) and Minter & Retkute (2019)). For ABC-SMC, we set the distance function as:

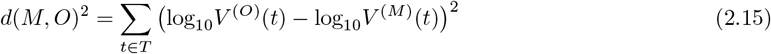

where the time points *t* are those in the set *T* where *V* ^(*O*)^ *> V*_*L*_ = and *V* ^(*O*)^ and *V* ^(*M*)^ represent the observations (from HCS) and simulated outputs respectively. We set 8 generations of iteration and collect 250 particles from iterations. During each iteration, we set the values calculated from the cost function smallest to largest and choose the value of the 1st quartile as the tolerance level for the next iteration.

In this work, the prior distribution for the replication phase parameters are given by:

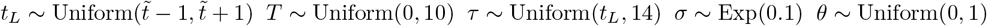

where 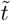 is the last day that viral load stays undetectable after deliberate infection. This means the time of peak must be after or same as the time of earliest detection In the supplementary material, Figures 5 and 6 show the posterior predictions (central estimate and 95% credible interval) and the observed data. Furthermore, in striving for more intuitive results, we merge the posterior distributions of each parameter assuming equal weighting to each sample, which is illustrated in Figure 1.

**Figure 1:**
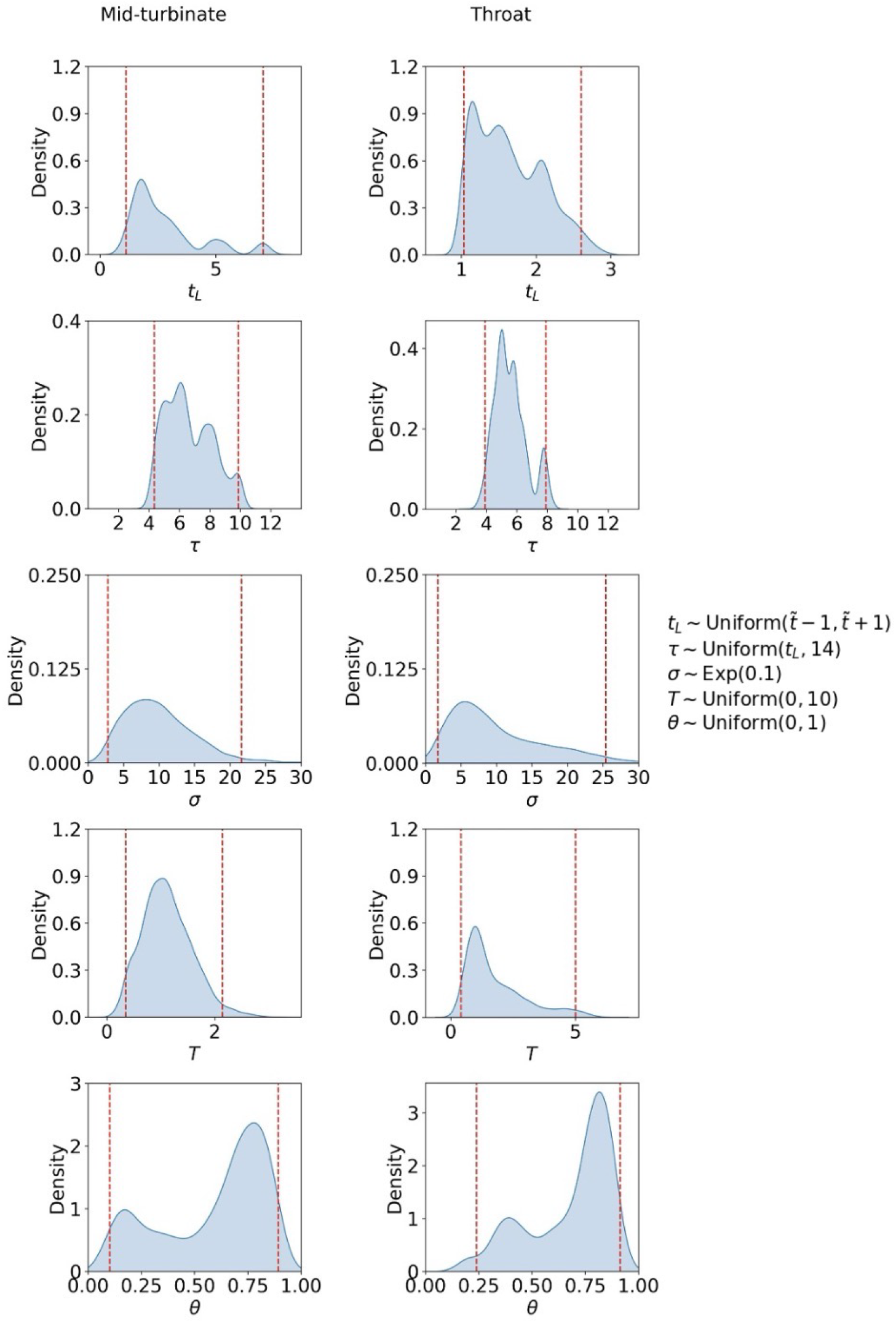
Posterior distribution of parameter values from the result of ABC-SMC for Model (2.5) using mid-turbinate data and throat data (Killingley et al. (2022)). The vertical dash lines represent the 2.5th and 97.5th percentiles for each parameter.

However, the adjustment model has two further parameters (*α* and *t*\*). We assume that for some *t*\* *< t*_*L*_ that *V* (*t*) = 0 for *t < t*\*. We may then match this to the adjustment phase solution such that 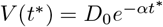 so for a choice of *t*\* we have

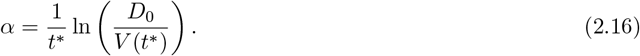

Then (2.14) enables us to obtain an estimate of *t*\* (given values of *D*_0_ and *P*_*I*_ (*D*_0_)) such that the following is satisfied

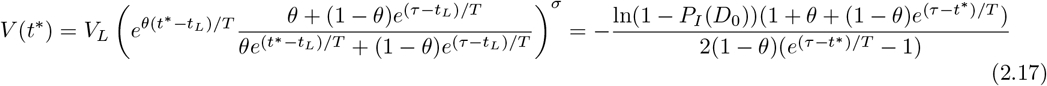

Note that *t*\* appears a number of times in (2.17) and so we may solve this numerically. However, an approximate estimate can be derived by making the assumption that *V* is growing exponentially and that *τ* ≫ *t*\* so that

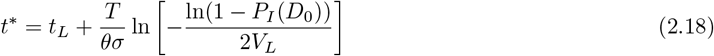

The Human Challenge Study tells us *D*_0_ = 55 and that 18 out of 34 volunteers developed an infection so *P*_*I*_ (*D*_0_) = 18*/*34 = 0.53 (Killingley et al. (2022)). The uncertainty in *α* and *t*\* is conditional on the posterior distributions elicited from data calibration.

## 3. Results

### 3.1. Model Calibration

We first calibrate the model (2.5) to the HCS and the merged posterior distributions for each of the parameters based on throat and mid-turbinate data are illustrated in Figure 1 (and the corresponding posterior predictions are illustrated in Supplementary material [Figures 5 and 6]). The red vertical lines on each panel show the 95% confidence interval on the merged posterior but as these are merged from a relatively small sample (18 volunteers) this should be treated as indicative only.

Note that the parameters *T* and *σ* appear fairly uni-modal and smooth for both sites, though the distribution of *T* is more narrow in mid-turbinate than throat. However, *t*_*L*_ and *τ* (both parameters sensitive to the location of the viral load in time) are more disjoint, due to the sample size of HCS, with the mid-turbinate having wider range than parameter estimates from throat data. The parameter *θ* appears bi-modal on both sites and generally with similar shape.

We can unpack these marginal summaries by looking at the posteriors distrbutions of each individual (Supplementary material Figure 7) but this is also seen in the bilateral correlation plots (Figure 2 and 3). It is worth reflecting on the action the parameters are having in the model: *t*_*L*_ and *τ* act to locate the viral load in time, *T* acts to scale the viral load over time (relative to *t*_*L*_ and *τ* ), *σ* scales the viral load magnitude while *θ* drives the shape. These plots also need a degree of care to interpret and are hard to briefly summarize, given that are a composite of 18 individual fits.

**Figure 2:**
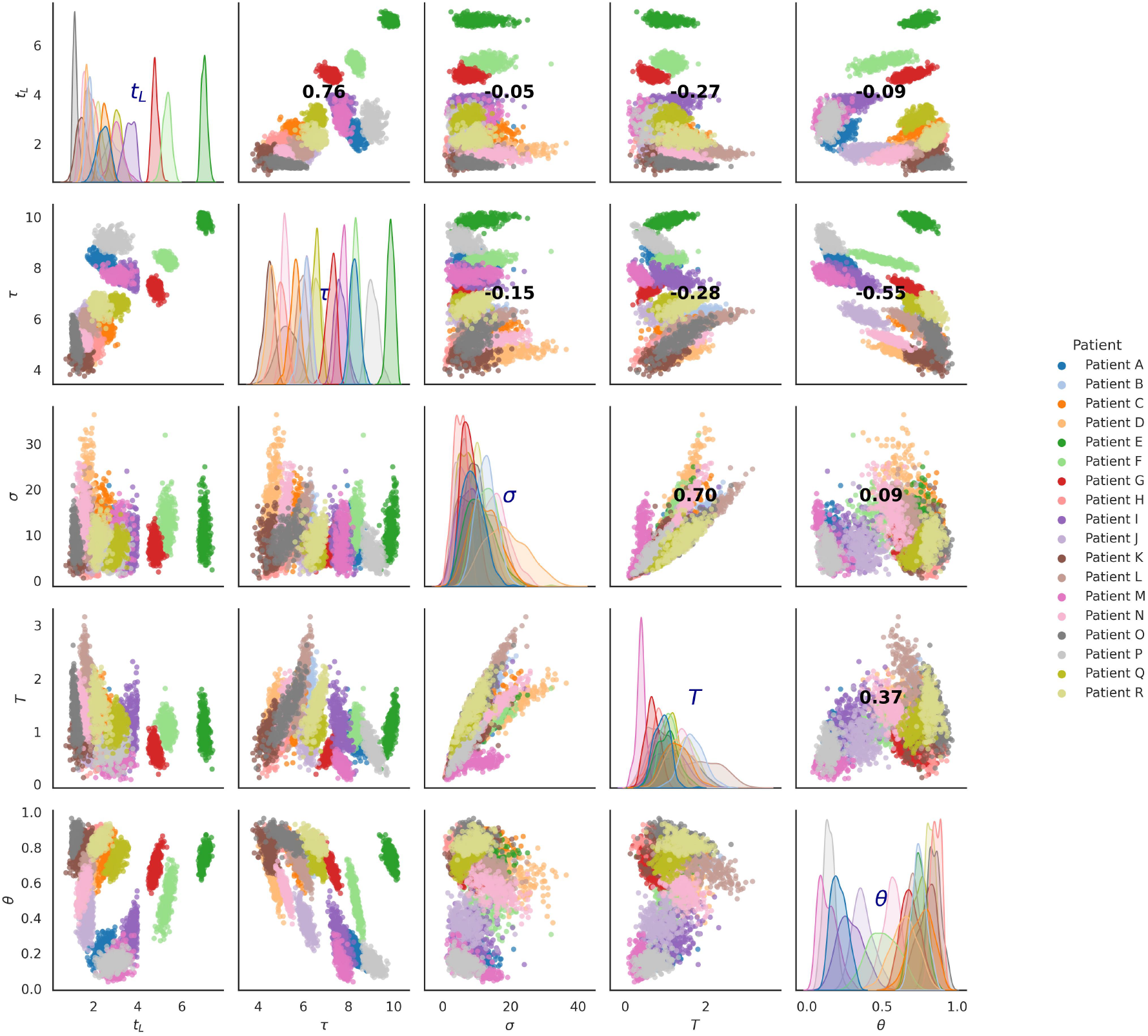
Scatter plots illustrating a pair plot showing the relationships among parameters from parameter inference based on mid-turbinate data (Killingley et al. (2022)). The diagonal plots show the distribution of each parameter, while the other plots illustrate the relationships between pairs of parameters. The bold numbers indicate the correlation strength between parameters.

**Figure 3:**
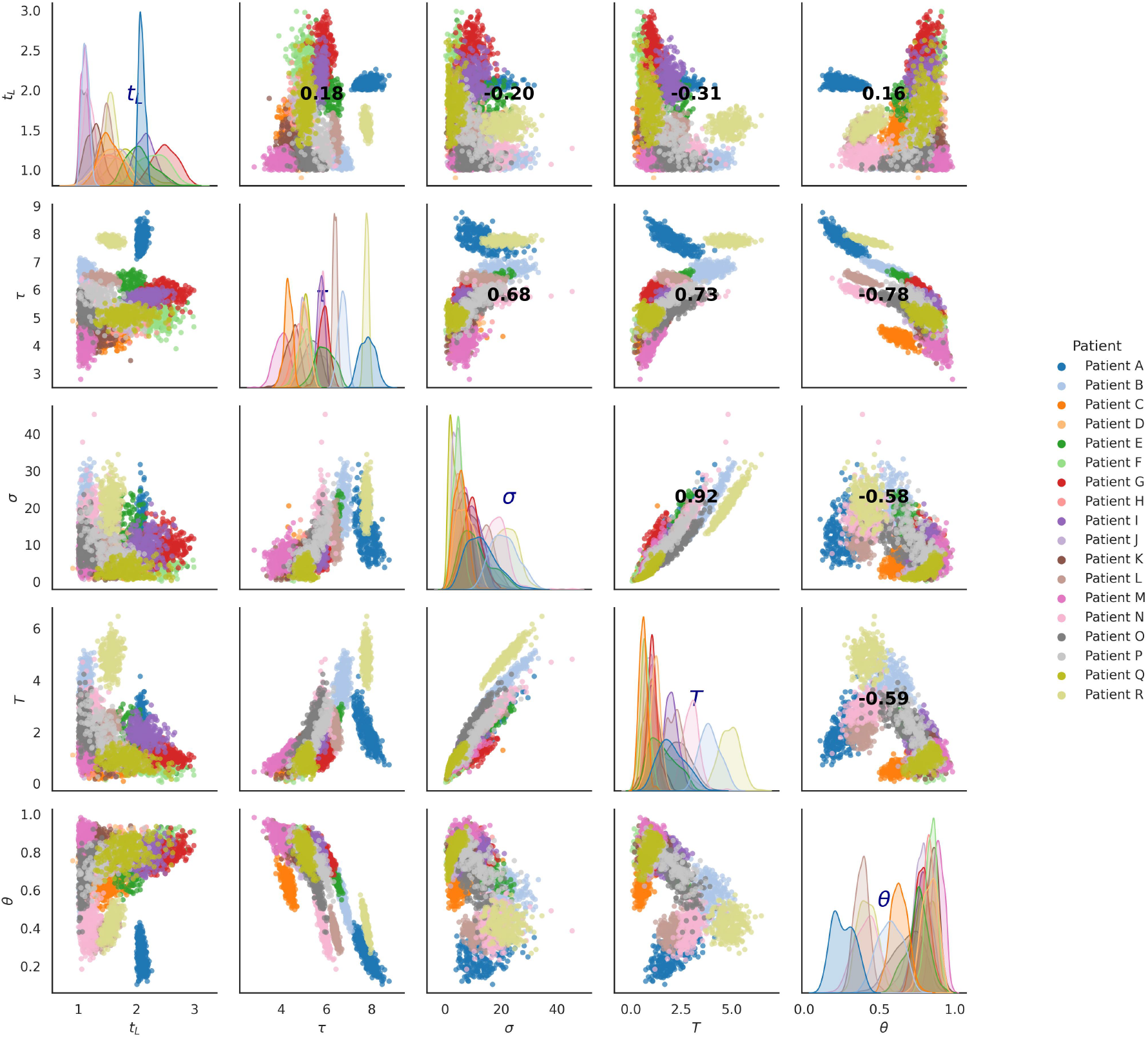
Scatter plots illustrating a pair plot showing the relationships among parameters from parameter inference based on throat data (Killingley et al. (2022)). The diagonal plots show the distribution of each parameter, while the other plots illustrate the relationships between pairs of parameters. The bold numbers indicate the correlation strength between parameters.

In Figure 2 the parameter estimates appear to show the strongest degree of overall correlation between *τ* and *t*_*L*_ (larger *τ* suggests larger *t*_*L*_) but looking at the individual parameter estimates by colour block this is less apparent (or may be negatively correlated). Figure 3 does not show the same pattern of overall correlation for *t*_*L*_ with other parameters.

The stronger patterns of correlation are a correlation between *T, τ* and *θ* in both locations. Not only are they consistent between sites but they also have broadly consistent trends between individuals. The timescale of immune response (*T*) is positively correlated with the exponent for viral load (*σ*) for individuals and across whole sample so larger values of *T* (slower immune response activation) correspond to larger values of *σ* (meaning potentially higher eventual viral concentration at peak). This does not mean that people with slower immune response necessarily shed more (*θ* and *τ* would have some role in this too).

The interplay between the timing of the peak *τ* and *θ* appears more strongly in the throat than mid-turbinate but is consistent in both (and correlations in general are weaker in mid-turbinate). Individuals appear to cluster in two groups, one with low *θ* and high *τ* and *visa versa*, with majority of individuals in latter. Although the individual scatter in Figures **??** and 3 shows a degree of complexity to this pattern this may be a tentative sign the people those viral load peaks later have slower growth rates. This may not sound particularly interesting as slow growth would suggest later peak but this is not strongly coupled with the magnitude of peak virus (driven by *σ*).

Based on the posterior distribution illustrated in Figure 1, we can easily estimate the value of *t*\* as well as the value of *V* (*t*\*) through equations (2.18) and (2.17) which are shown in Figure 4. Clearly we could have chosen to plot estimate for *α* instead of *V* (*t*\*) but felt that *V* (*t*\*) was more intuitive. The majority of individuals then have an adjustment time between 0 and 3 days in both locations though about 3 individuals have longer adjustment time in mid-turbinate (about 5 or 7 days). This is not surprising due to the values of *t*_*L*_ derived in previous section. However, the residual viral load at this time tends to be lower in mid-turbinate.

**Figure 4:**
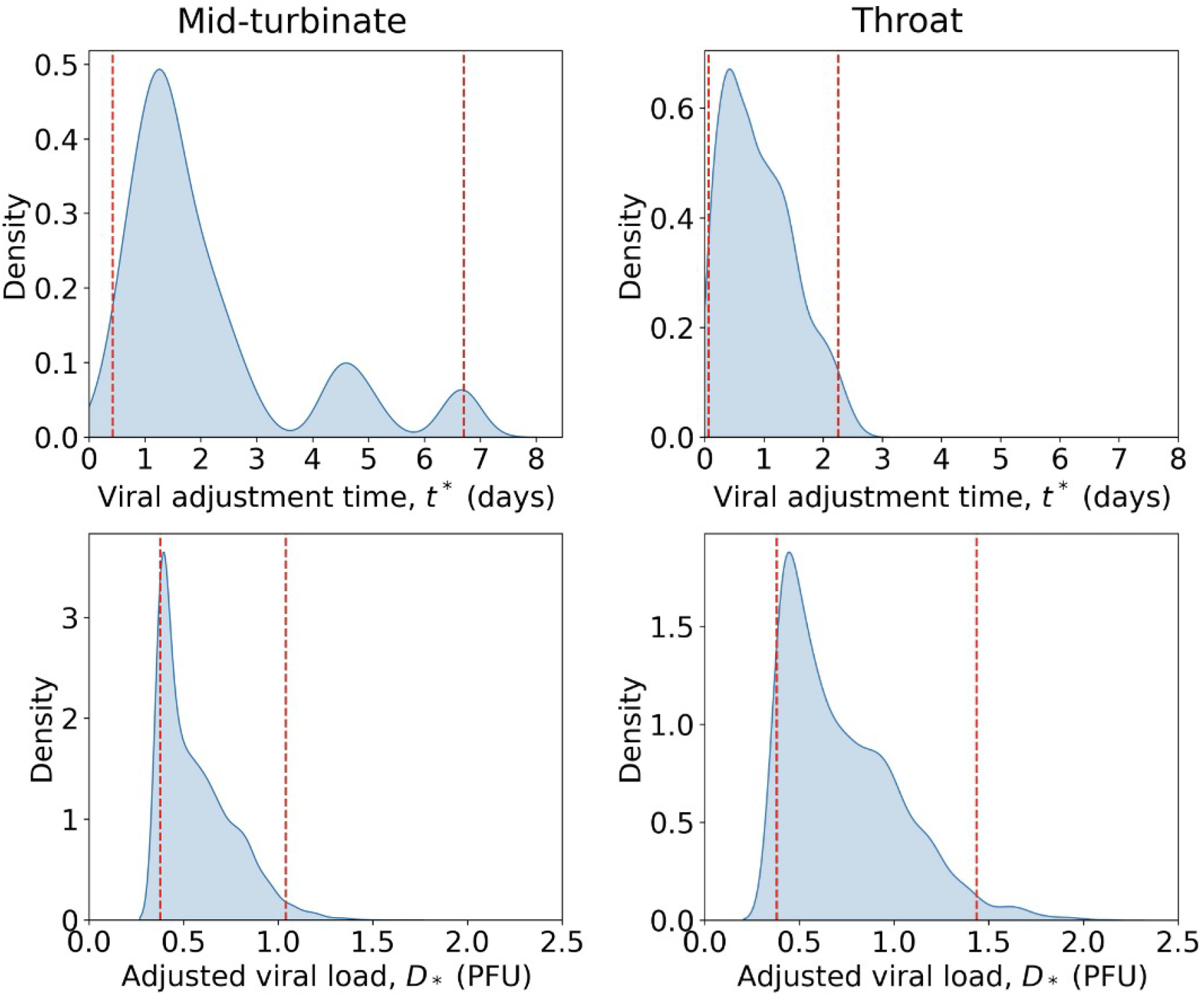
The first column: approximate posterior distributions of viral adjustment time *t*\* and adjusted viral load *V* (*t*\*) based on mid-turbinate data (Killingley et al. (2022)). The Second column: approximate posterior distributions of viral adjustment time *t*\* and adjusted viral load *V* (*t*\*) based on throat data (Killingley et al. (2022)). Other parameter inference results can be seen in Figure 1. The vertical dash lines represent the 2.5th and 97.5th percentiles for each parameter.

Apart from the difference in adjustment period, the adjusted viral load in mid-turbinate and throat show differences. In the left bottom of Figure 4, the kernel density estimation of *V* (*t*\*) of mid-turbinate shows that the actual viral load adjusted is around 0.5 to 1.6 PFU based on mid-turbinate data, which is a loss of 2 orders of magnitude from the initial dose, 55PFU. For mid-turbinate, the adjustment rate is around 1%-3%. In the right bottom of Figure 4, the adjusted viral load is around 0.5 to 2.5 PFU. This result could be indication that throat may be more susceptible to SARS-CoV-2 virus and more virus can survive viral adjustment or may be an artefact of the dosing regime of HCS. Overall, Figure 4 indicates that a very small amount of virus retained in human body is enough to cause an infection.

### 3.2. Simulation of the stochastic viral adjustment phase

Due to the difficulty of finding the analytical solution for the probability of clearance during viral adjustment phase, we will carry out simulation to see the viral dynamics. The idea is that we first simulate the SDE (2.2) for *t* ∈ [0, *t*\*]. If *D*_*t*_ hits 0, we consider the virus is cleared before virus enters susceptible cells and the patient will not develop an infection. Otherwise, *V* (*t*\*) *>* 0 gives the adjusted viral load at *t*\*. For each patient, we use the mean value of the posterior distribution for each parameters, *t*_*L*_, *T* , *τ* , *θ* and *σ*, from which we can easily calculate the value of *α* and *t*\* through Equations (2.16) and (2.18). For each patient, we simulate the corresponding SDE (2.2) for 30,000 times, from which we can receive the proportion of simulation that faded out during viral adjustment. Since we know that the Human Challenge Study indicates the overall viral clearance rate is 0.47, multiplying 0.47 with the proportion of clearance during viral adjustment will enable us to know the probability of clearance during viral adjustment and viral replication respectively. The results of our simulation are displayed in Table 1 and also in supplementary material Figures 8 and 9.

**Table 1:** Simulated values for viral adjustment time *t*\*, viral decay rate *α*, fade out rate in adjustment, and Clearance rate in viral replication based on the posterior distribution in Figure 1.

| Patient No. | $t^*$ | $\alpha$ | Fade out probability during adjustment |
| --- | --- | --- | --- |
| A | 1.13 | 4.40 | 0.68 |
| B | 1.59 | 3.08 | 0.66 |
| C | 2.74 | 1.68 | 0.55 |
| D | 1.82 | 2.59 | 0.60 |
| E | 6.67 | 0.70 | 0.57 |
| F | 4.55 | 1.07 | 0.64 |
| G | 4.96 | 0.93 | 0.55 |
| H | 1.54 | 3.04 | 0.60 |
| I | 2.18 | 2.27 | 0.66 |
| J | 1.01 | 4.87 | 0.66 |
| K | 0.96 | 4.88 | 0.60 |
| L | 1.39 | 3.48 | 0.64 |
| M | 2.44 | 2.02 | 0.68 |
| N | 1.02 | 4.79 | 0.66 |
| O | 0.66 | 6.45 | 0.43 |
| P | 1.10 | 4.53 | 0.68 |
| Q | 2.59 | 1.84 | 0.62 |
| R | 1.78 | 2.62 | 0.57 |

Table 1: Simulated values for viral adjustment time $t^*$ , viral decay rate $\alpha$ , fade out rate in adjustment, and Clearance rate in viral replication based on the posterior distribution in Figure 1.
| Patient No. | $t^*$ | $\alpha$ | Fade out probability during adjustment |
| --- | --- | --- | --- |
| A | 0.29 | 12.45 | 0.64 |
| B | 0.76 | 5.36 | 0.34 |
| C | 1.24 | 3.71 | 0.55 |
| D | 0.76 | 6.19 | 0.60 |
| E | 1.36 | 3.57 | 0.64 |
| F | 1.66 | 2.80 | 0.57 |
| G | 1.74 | 2.80 | 0.64 |
| H | 1.34 | 3.45 | 0.55 |
| I | 0.78 | 6.17 | 0.64 |
| J | 1.48 | 3.16 | 0.57 |
| K | 0.29 | 15.83 | 0.53 |
| L | 0.43 | 10.80 | 0.55 |
| M | 0.77 | 4.97 | 0.23 |
| N | 0.89 | 4.61 | 0.36 |
| O | 0.92 | 4.10 | 0.21 |
| P | 1.44 | 3.25 | 0.57 |
| Q | 1.65 | 2.63 | 0.45 |
| R | 0.32 | 12.75 | 0.36 |

In Table 1, both mid-turbinate and throat show similar fade out probabilities in viral adjustment with mean 0.29 and 0.26 respectively. This means about 60% of clearance is achieved during the adjustment phase. For example patient A has 32% chance of not becoming a case due to the adjustment phase, then they had a residual viral load at start of replication phase that carried a 22% chance of stochastic fade out (conditional on virus surviving to replication and the overall 47% infection rate in volunteers).

## 4. Discussion

In this paper, we develop explicit mechanisms to explain viral dynamics in the first few days of infection *and* evaluate the probability of infection by deriving the dose-response model. Furthermore we extend the viral load model by considering logistic growth in adaptive immune response. We fully appreciate these models are coarse simplifications of the true biological mechanisms but they enable us to interpret the available data.

Based on Figures 2 and 4, we notice that the mid-turbinate overall shows a broader window for viral adjustment: the virus will achieve viral adjustment within three days for nearly half of the patients. However, there are patients show much longer period of adjustment phase ranging from 0 to 8 days, which is unsurprisingly consistent with the longer undetectable period in some patients (cf. Figure 5 in Appendix). Furthermore, the throat cases show a narrower length of adjustment phase up to 3 days. For this phenomenon, we consider there are a few explanations. First, the virus will infect in throat first and replicate during the early phase of infection and then the virus re-migrates to mid-turbinate to pose this broader period of adjustment. Secondly, one considers that the throat possesses more susceptible cells for SARS-CoV-2 than mid-turbinate. Another hypothesis could be that the susceptible cells in mid-turbinate have higher level resistance to SARS-CoV-2. Based on our current data and knowledge, we cannot identify which explanation is more likely.

The stochastic viral adjustment phase allows virus to get completely cleared before it enters susceptible cells. This viral clearence can be caused by immune response or failure to deposit in suitable site. As we have used the Human Challenge Study dose response observation that 47% of cases never have detectable virus, the stochastic viral adjustment mechanism reflects this strong chance that the virus will be cleared in either the viral adjustment or replication phases but suggests that in about 2/3rds of those that are not infected following exposure this is due to adjustment phase mechanisms. Of course what we are calling the adjustment phase is much more omplicated and may involve many potential bottlenecks or decay rates for which our model of 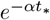 is an crude approximation.

However, based on Table 1, we can see that throat shows overall shorter *t*\* than that of mid-turbinate, which is consistent with what we observed in the deterministic viral adjustment case. As we mentioned above, the throat cells may be more susceptible so the viral adjustment phase is shorter. Also, the values of *α* of mid-turbinate has mean 3.07, and this is smaller than the value of *α* in throat which is 6.03. This difference in *α* indicates that the throat may have stronger immune response, which clears the virus faster, or people have high chance to clear in their throat which reduces the viral load compared with say sneezing. Figures 8 and 9 in supplementary materials illustrated the distribution of viral load that adjusted to the body, in which we can see that in general, both mid-turbinate and throat have similar range of values. However, mid-turbinate has a predominantly low viral load with limited variation, while throat has a more varied and higher range of adjusted viral load values, which is consistent with the previous conclusion of different viral dynamics in mid-turbinate and throat. It should be pointed out that in Figure 8 and 9, the value only means the viral load that enters the viral replication phase and does not mean the viral load can guarantee the infection, which should be distinguished from the deterministic case.

In this paper, we constructed a boundary problem based on the viral process, which leads to a dose-response model with exponential form. Upon our best knowledge, boundary problem has not previously been used to develop dose-response models and in this case returns the form expected from competing risk arguments but may allow for greater flexibility with further research. Under the competing risk framework of bacteria infection, Haas et al. (2014) develops a series of models to investigate dose-response relationship in different cases ranging from theoretical experimental scenarios to complicated real-world scenarios. In Haas et al. (2014) (also cf.Xu et al. (2023)), the assume that the virus is randomly distribution in the atmosphere around the inhaler and expected dose will be proportional to the amount of inhaled air. Whilst of similar mathematical form, the competing risk derivation is designed for pooled data so assumes that all patients are homogeneous given a specific dose while Model (2.14) enables consideration the heterogeneity of individuals based on each set of the viral load data.

The viral replication model proposed in this study explains the data as well as conventional SIV-type models or models with more structured models for adaptive immune response. This model has 5 parameters, two are directly intuitive: time of detection and time of peak (*t*_*L*_ and *τ* respectively). The others (*θ, T* and *σ*) have an interpretation from model derivation in terms of the relative differences between timescales but they can also be considered as functions of the growth rate (i.e. *θσ/T* ), decay rate (i.e. (1 − *θ*)*σ/T* ) and peak viral load, see Supplementary section **??**. This means the parameters are identifiable provided supporting data grows and then decays.

The striking finding is that the parameter *θ* shows signs of bimodality in the sample of volunteers. This is a clear feature of this sample but it is not clear if this is an artifact of a relatively small sample (so more volunteers would have produced a more unimodal posterior combined sample) or a genuine feature. The estimates for *t*_*L*_ and *τ* appear to be multi-modal but as a result of the sample size of volunteers and that more participants might be expeted to smooth these merged posteriors out further. However, the shape of posterior for *θ* appears structurally different particularly in mid turbinate sample.

The parameter *θ* is the the growth rate divided by the sum of growth and decay rates (*θ* = *r/*(*r* + *r*_*D*_)). So slow growth with fast decay would lead to smaller values of *θ*, fast growth and slow decay values closer to 1 and balanced growth and decay values close to 0.5. From Figure 2, in the mid-turbinate, those samples with smaller *θ* appear to exhibit later times of peak (*τ* ) as might be expected for relatively slow growth (and consistent values of *t*_*L*_). However, those with faster growth than decay exhibit a wider range of peak and detection times. It is noteworthy too that those volunteers with the highest eventual viral load in mid-turbinate (J and M) are in this cluster. This pattern is less clear in throat data.

## Data Availability

All data used in the present study are publicly available.

## Funding information

JX, MLG, TH, and IH were supported by the TRACK: Transport Risk Assessment for COVID Knowledge project - EPSRC, EP/V032658/1 -.

## Acknowledgment

JX, MLG, TH, and IH were supported by the TRACK: Transport Risk Assessment for COVID Knowledge project - EPSRC, EP/V032658/1 -. The authors would like to thank the rest of the TRACK team.

Ian Hall

School of Mathematics

The University of Manchester

Oxford Road

Manchester M13 9PL

United Kingdom

## Supplementary Material

### A. Continuous time Markov chain for viral adjustment phase

In the previous section, we add scaled noise to model (2.1) to obtain the stochastic viral adjustment phase. However, the SDE (2.2) does not give an analytical solution. Hence, we can construct a continuous time Markov chain, *D*_*t*_ ∈ [0, ∞) for *t* ∈ [0, ∞), for the viral adjustment phase. The distribution of *D*_*t*_ at a given point of time *t* can be easily calculated.

We assume that the lifespan of each virus is exponentially distributed with rate *α* ∈ (0, ∞), and each virus is independent. In this case, we see that, for *t*∈ [0, ∞ ), the probability of a single virus survived after *t* is:

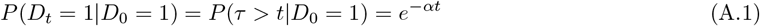

where *τ* is the survival time that has the exponential distribution with parameter *α >* 0. Hence, for initial dose *D*_0_, the distribution of *V* (*t*\*) when the virus achieves viral adjustment at *t*\* follows a binomial distribution with success probability 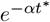.

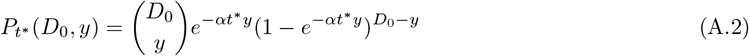

where *y*∈ { 0, 1, …, *D*_0_}. Note that one limitation of the CTMC is that it only allows integer values of *y*. The probability of survival in A.1 is equivalent to the probability of realising viral adjustment. Hence, based on the posterior distributions in Figure 1 and the equations (2.16) and (2.18), we can easily estimate the value of 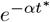. In figures 10 and 11, we sample 100,000 times based on the distribution A.2. We can see that figures 10 and 11 lead to similar conclusion as Figures 8 and 9.

### B. Supporting Figures

**Figure 5:**
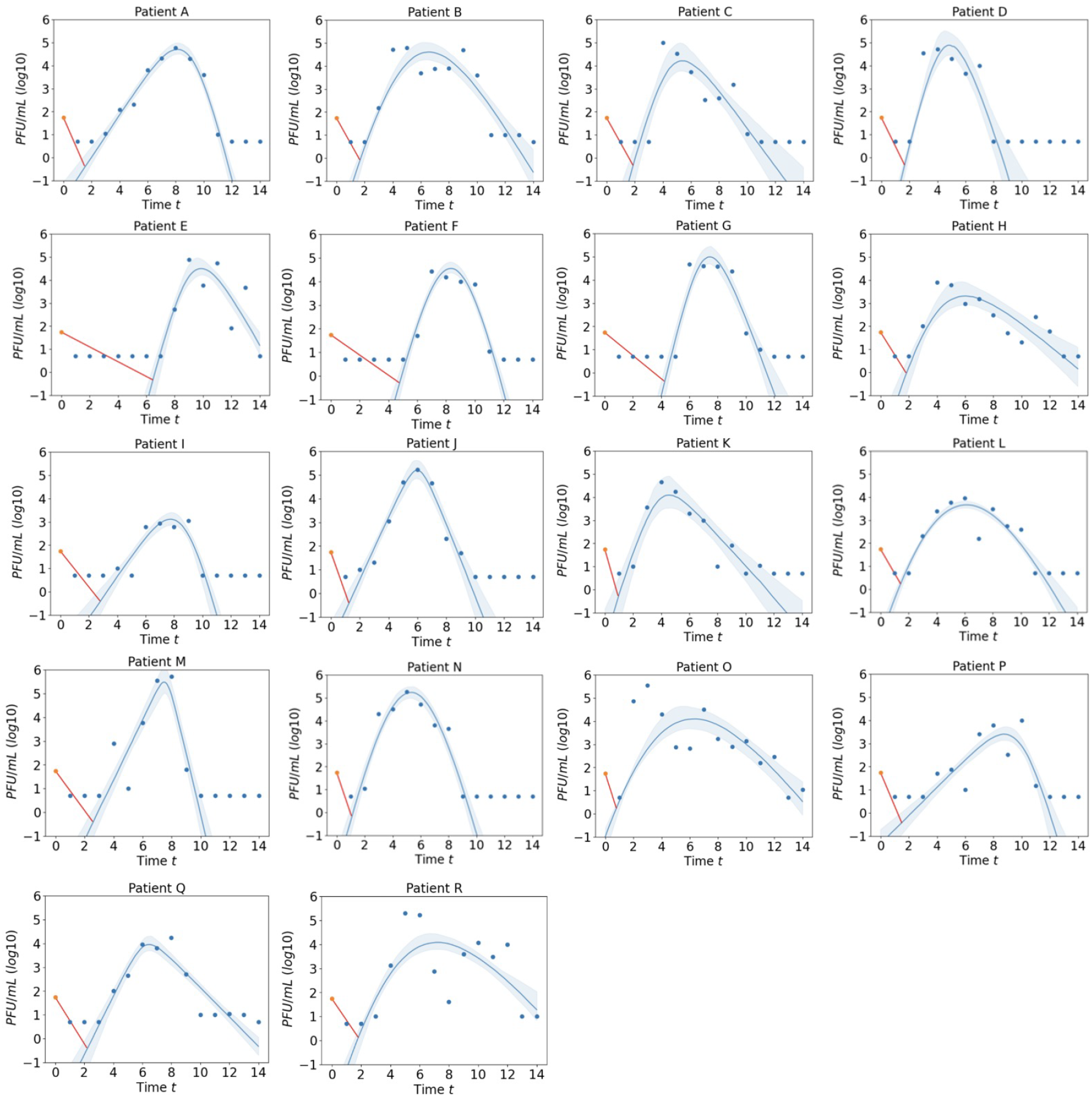
Posterior predictions of infectious virus (PFU/mL) from mid-turbinate for 18 participants from the Human Challenge Study (Killingley et al. (2022)). Model is that derived in equation (2.5). Shaded regions represent the 95% credible interval. The orange dot at day 0 represents the initial dose, which is 55PFU/ml and the red trajectory represents the viral decay during the viral adjustment phase.

**Figure 6:**
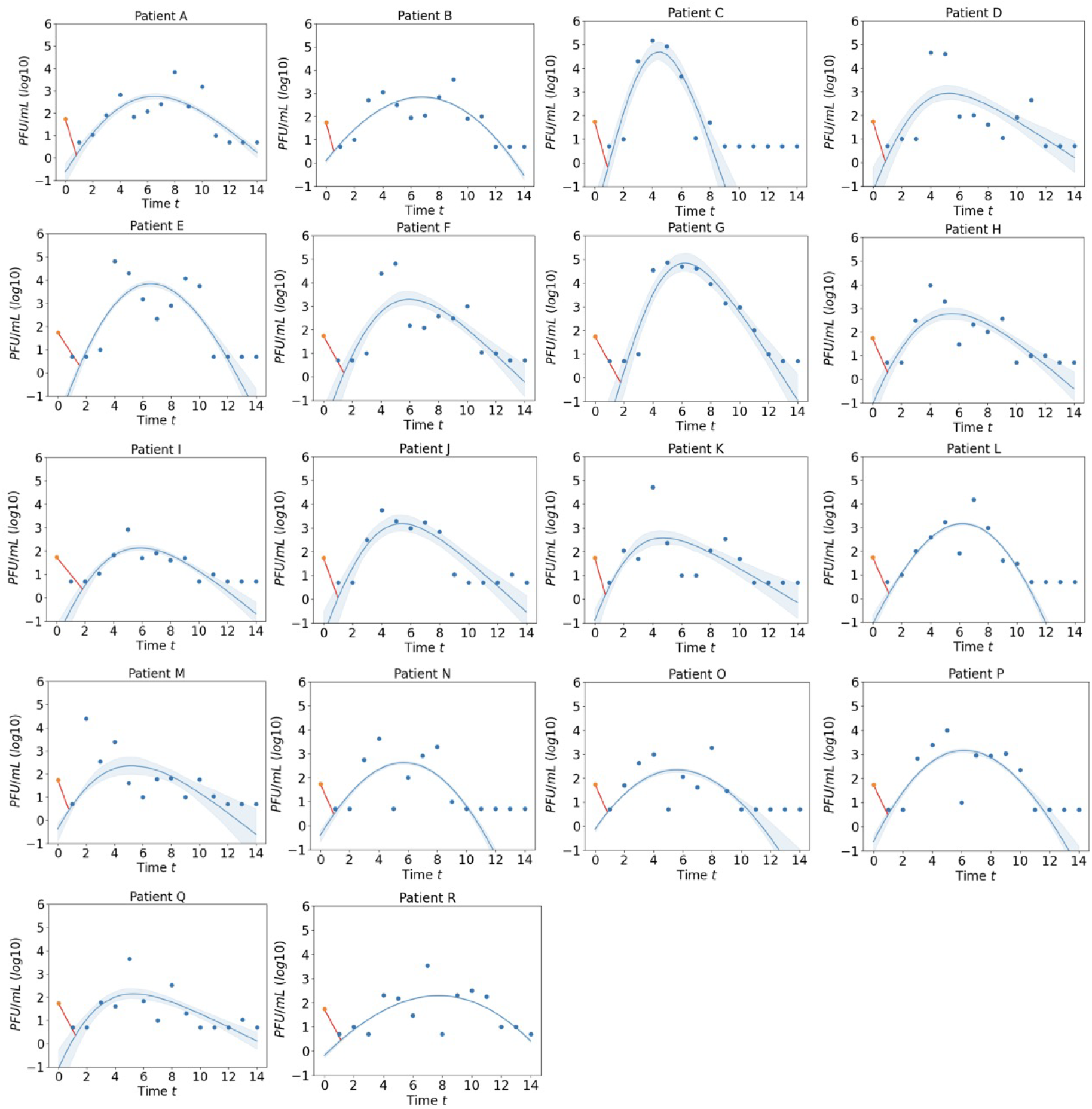
Posterior predictions of infectious virus (PFU/mL) from throat for 18 participants from the Human Challenge Study (Killingley et al. (2022)). Model is that derived in equation (2.5). Shaded regions represent the 95% credible interval. The orange dot at day 0 represents the initial dose, which is 55PFU/ml and the red trajectory represents the viral decay during the viral adjustment phase.

**Figure 7:**
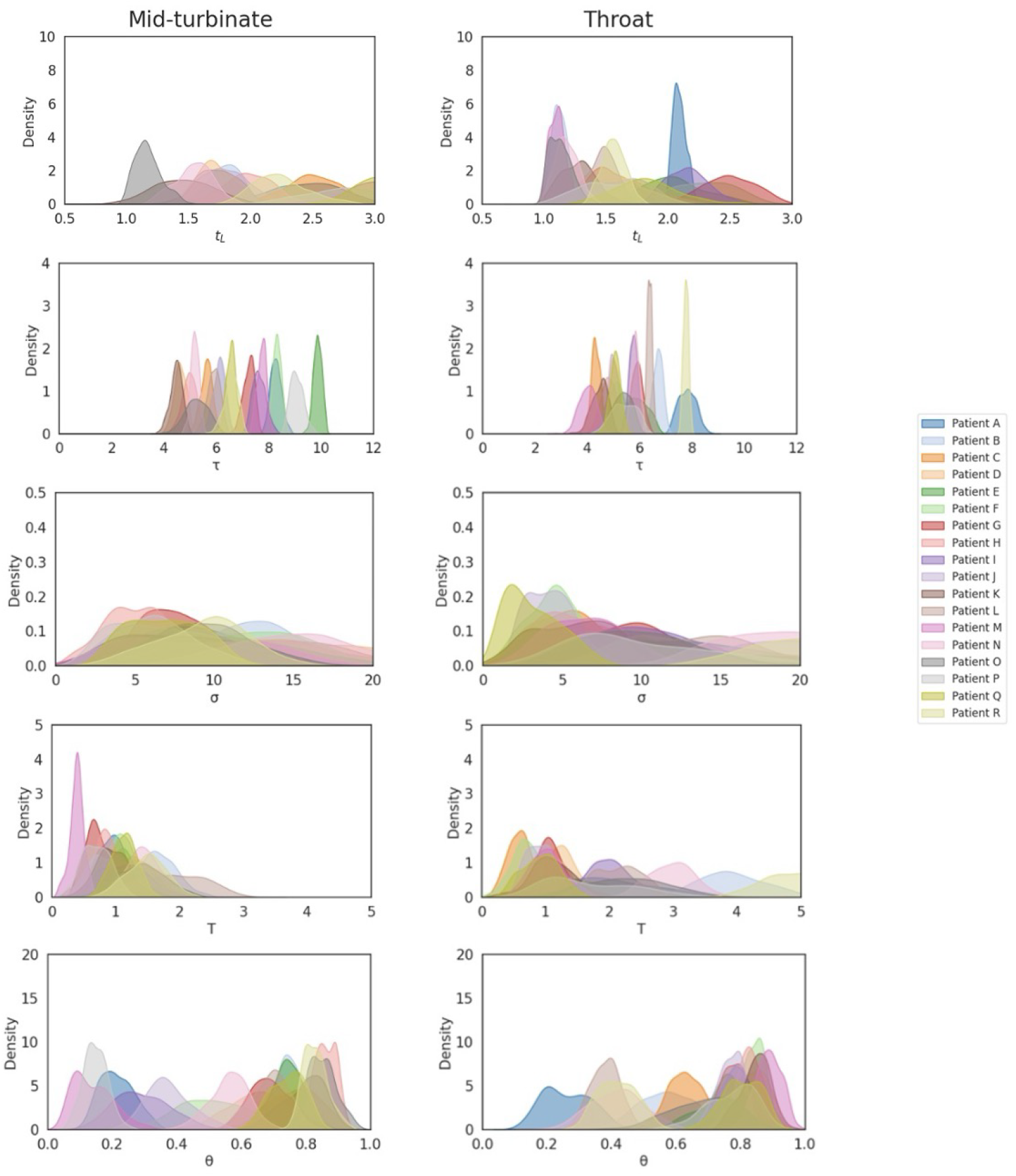
Posterior distribution of parameter values from the result of ABC-SMC with model from Model (2.5) using mid-turbinate data and throat data (Killingley et al. (2022)).

**Figure 8:**
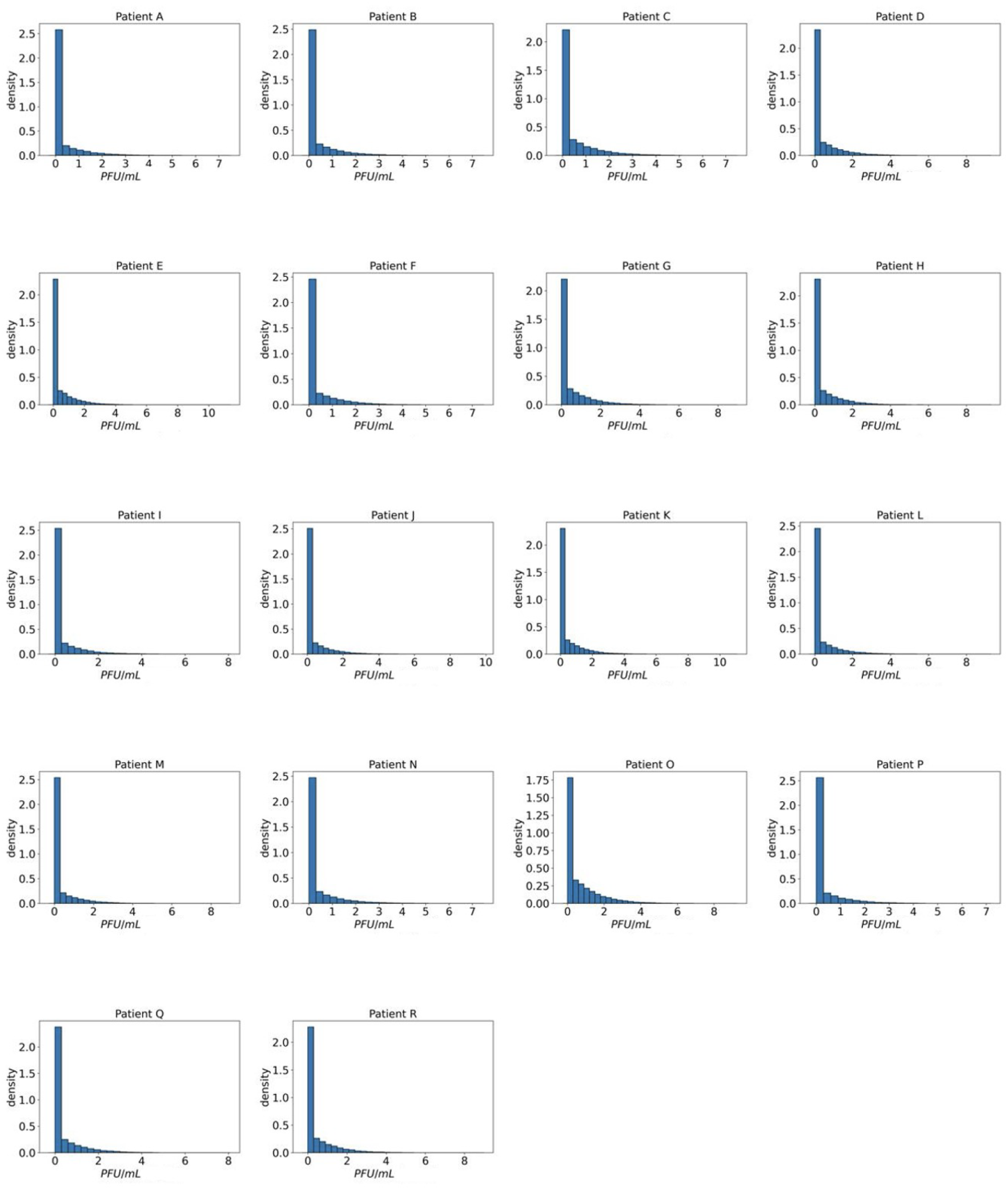
Distribution of simulated viral load that successfully enter susceptible cells, *D*\*, based on mid-turbinate data

**Figure 9:**
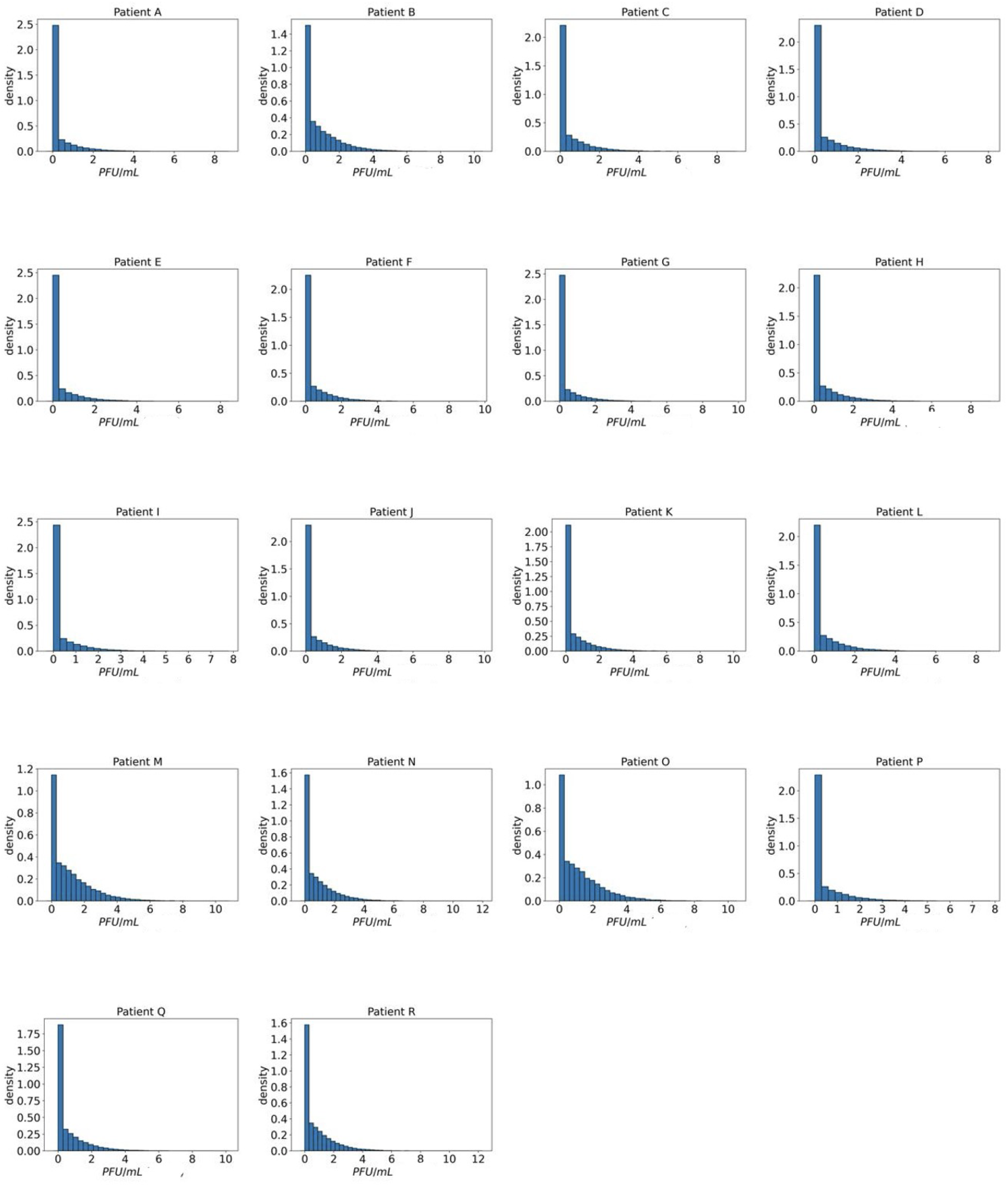
Distribution of simulated viral load that successfully enter susceptible cells, *D*_*_, based on throat data

**Figure 10:**
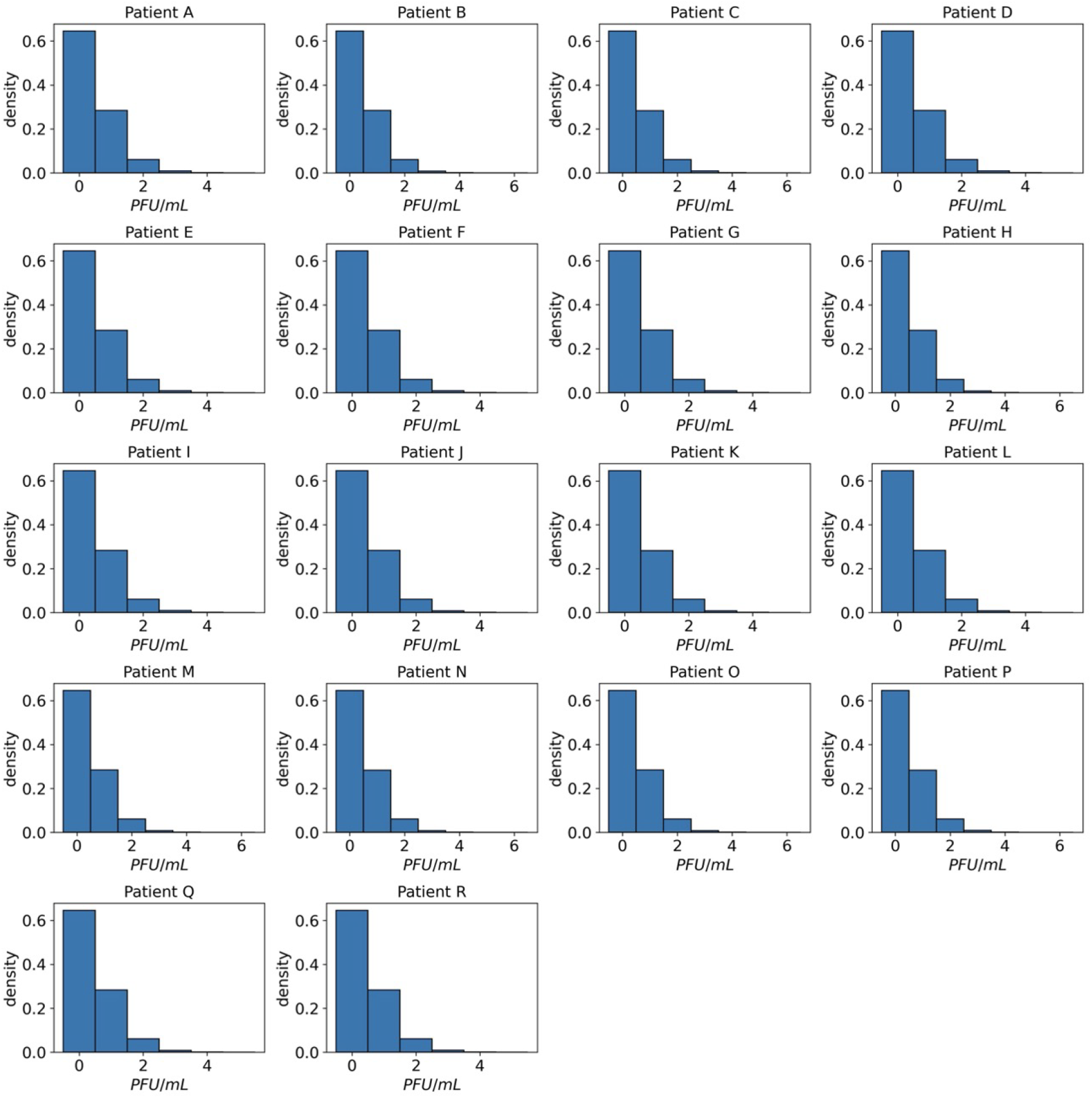
Continuous time Markov Chain case: distribution of simulated viral load that successfully enter susceptible cells, *D*_*_, based on mid-turbinate data (Killingley et al. (2022)). The x-axis represents the viral load in PFU/ml

**Figure 11:**
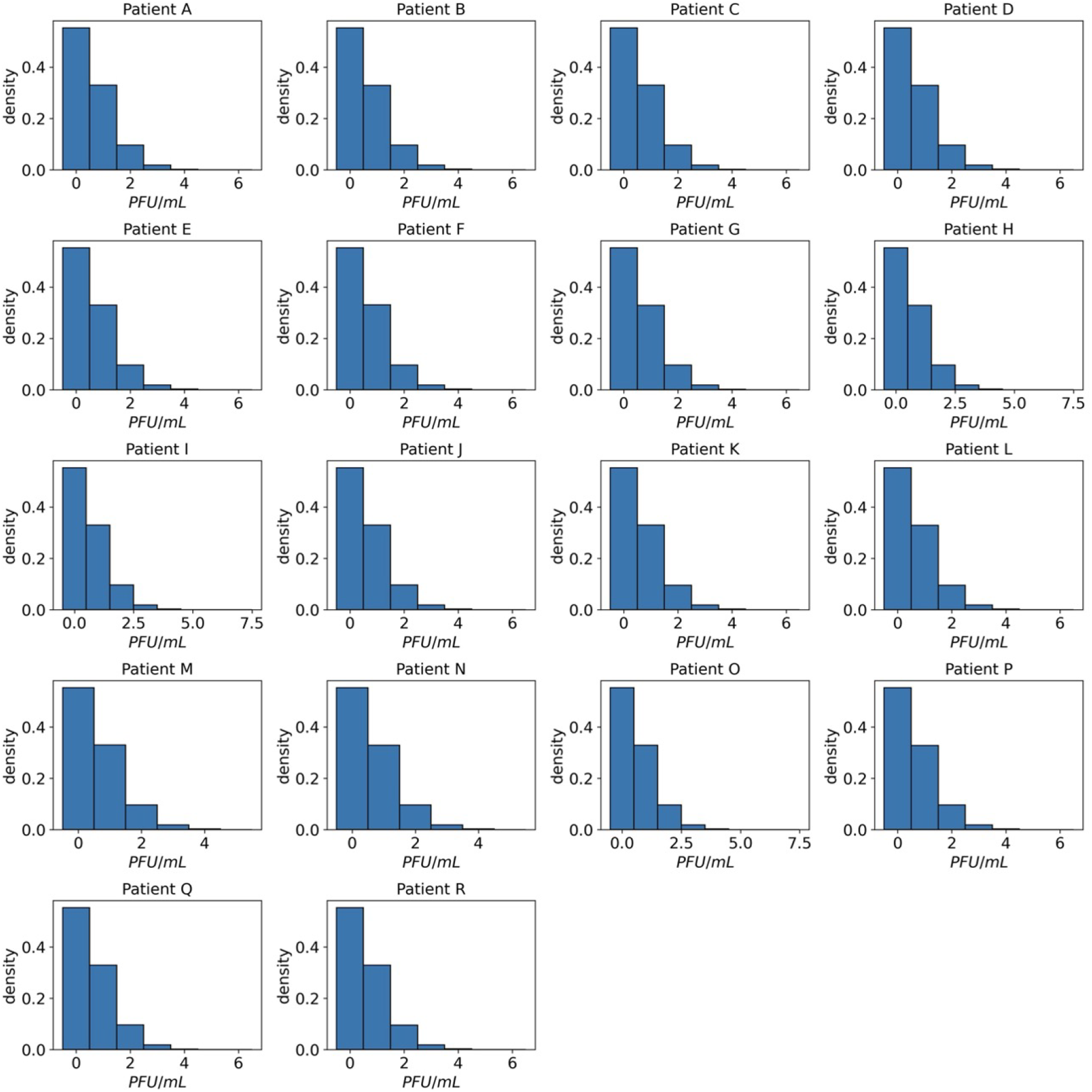
Continuous time Markov Chain case: distribution of simulated viral load that successfully enter susceptible cells, *D*_*_, based on throat data (Killingley et al. (2022))

## Notes

### Competing Interest Statement

The authors have declared no competing interest.

### Funding Statement

all authors were supported by the TRACK: Transport Risk Assessment for COVID Knowledge project - EPSRC, EP/V032658/1 -

### Author Declarations

Killingley et al. (2022), Safety, tolerability and viral kinetics during sars-cov-2 human challenge in young adults, Nature Medicine. https://www.nature.com/articles/s41591-022-01780-9

